# Protocol for a qualitative study exploring older adults’ experience of mental health and wellbeing in work-related later life transitions

**DOI:** 10.1101/2025.01.23.25321020

**Authors:** Rebecca Woodhouse, Dean McMillan, Ruth Wadman

## Abstract

Later life transitions occur in the later years of life and can cause significant disruption to a person’s usual daily routine and lifestyle. Transitions that are specifically related to work, such as retirement, can cause sudden changes to an individual’s financial, social and psychological environment and for some, may negatively impact on mental health and wellbeing. This protocol outlines a qualitative study to explore the experiences of adults who have experienced a negative impact on their mental health or wellbeing due to a work-related transition in later life. Semi-structed interviews will be completed with people aged 50 and over to gain a deeper understanding of 1) the mental health and wellbeing of people undertaking later life transitions; 2) the context surrounding the transitions (such as due to ill-health or redundancy); and 3) the types of support they received or may have benefitted from. Research in this area is scarce and this study will help us to better understand the experience of those who have encountered such difficulties to help develop appropriate interventions and support in this area.

## Introduction

Later life transitions are transitions that occur in the later years of life and can have a significant impact on a person’s lifestyle or environment. As these types of transitions create such significant changes, they can sometimes have a negative impact on mental health and wellbeing (Dang et al. 2022). This study is focussed on transitions that are related to work, such as retirement, role or career change, a reduction in hours from full to part time, moving to a type of ‘bridge-employment’ (paid work undertaken after main career job, but before full retirement) or volunteering.

Retirement is the transition that tends to be most frequently reported on. It can force a wave of changes on an individual, that may be sudden and somewhat unexpected, impacting both mentally and physically (Dave et al. 2008). Transitioning from employment to retirement often leads to the removal or reduction in financial security, fewer opportunities for socialisation, losing a sense of accomplishment and a regular routine; often factors in keeping well (Dang et al. 2022). Retirement and other work-related transitions are experienced differently for everyone and the contexts that surround them are also widely varied (Dave et al. 2008; Scharn et al. 2018). Contexts surrounding these transitions and the adjustment that follows can include time to prepare, personal choice, and social circumstances (Liu et al. 2019).

There are findings reporting both positive and negative experiences for later life transitions, specifically for retirement. A recent review reported a third of retirees had depression during their retirement transition (Pabon-Carrasco et al. 2020), whist another has shown retirement can be beneficial by improving depressive symptoms (van der Heide et al. 2013) or reducing the risk of depression by nearly 20% (Odone et al. 2021). Various factors may be able to account for some of the variations, for example, those forced to undertake a transition, such as involuntary retirement due to redundancy or ill-health, led to poorer health outcomes compared to those not forced to retire (Donahue et al. 2006). In addition, the majority of existing studies are not focussed on the transition period itself with inconsistent time periods since retirement reported, with some exploring wellbeing in retirement with participants over 10 years since the actual transition.

Existing research has identified a number of factors that have impacted on a person’s daily life and wellbeing during retirement. One that is often reported is the changes experienced to person’s identity and role, which have a strong impact on a person’s daily life once they are retired (Cahill et al. 2021; Hobbis et al. 2011). Social interactions and relationships were of also noted to be of value, with an important need for emotional adjustment following retirement (Bauger et al. 2016; Cahill et al. 2021; Fadeeva et al. 2020; Genoe et al. 2022; Haigh et al. 2005; Hobbis et al. 2011; Pepin et al. 2011). These findings are important, to highlight the particular values and factors that are disrupted during later life transitions; though the studies are rather varied in their reporting periods (time since the transition) and are predominantly samples of retirees within general populations, not specifically those who have experienced a negative impact on mental health or wellbeing.

The lived experience of people undertaking work-related transitions in later life has been sparsely reported, especially for non-retirement work-related transitions such as late life career or role change. For retirement, existing qualitative studies have focussed on general populations of retirees rather than those who have specifically experienced a negative impact on their mental health or psychological wellbeing due to the transition. There is little (if any) published research focussing on the experiences of those who have experienced a negative impact on their mental health or psychological wellbeing in work-related later life transitions.

As research has reported, some people may struggle with their mental health and psychological wellbeing during or following a work-related transition in later life. It is therefore important to understand the experiences of those who have encountered such difficulties to develop appropriate interventions and support in this area.

## Materials and Methods

Aims: This study aims to explore the experiences of people who have experienced a negative impact on their mental health or psychological wellbeing due to undertaking a work-related transition in later life. The main areas of focus will be the experience and context of the transition and exploring the impact of the transition on mental health and wellbeing. The support and resources used by individuals will also be explored to identify factors that may have helped or hindered the transition period. In addition, the interviews will gather people’s perspectives on what help or support would have been acceptable to them during the transition: when the most beneficial timing to receive support would be; what that support may look like or involve; and who might provide it. This information will help to shape the future development of interventions to support people through work-related later life transitions.

### Research objectives

1. To explore the views and experiences of people who have experienced a negative impact on their mental health or wellbeing due to undertaking a work-related transition in later life
2. To explore the support and resource use of people undertaking work-related later life transitions
3. To identify acceptable supports and resources for work-related later life transitions to inform future research and the development of suitable interventions to support these transitions.

### Study design

This is an exploratory qualitative study using individual semi-structured interviews to explore the experiences and views of older adults who have recently undertaken a work-related transition. This type of interview was deemed to be the most appropriate due to the potentially wide variations in transitions, employment type and role, which may make group discussions difficult due to the uniqueness of everyone’s transition context. In addition, due to the sensitive nature of the discussions on mental health and wellbeing, focus groups were not chosen as they can limit people’s willingness to be open about their personal experiences without feeling embarrassed or feeling judgement, potentially limiting the depth of the results (Kitzinger, 1995). Though focus groups may have been fruitful in encouraging some discussion and offering different perspectives, they are particularly useful when studying a particular culture or group (Kitzinger 1995). Individual interviews can be easily conducted remotely, via telephone or video call, making it possible to include participants across a wide geographical region.

### Participants

Participants will be aged 50 and over and will have undertaken a work-related transition in the past five years, which has negatively impacted on their mental health or wellbeing. The transition will be related to work or employment, such as retirement, role or career change, or move to part time work or volunteering. The mental health and wellbeing difficulties will be self-identified by participants and not assessed using a standardised diagnostic measure.

### Inclusion criteria

- Adults aged 50 and over
- Undertaken a work-related transition in the past five years
- Experienced negative impact on their mental health or wellbeing around the same period as the LLT (self-reported).
- Living in the UK

### Exclusion criteria

- Unable to read and speak English.

### Sample size

A minimum of ten and maximum of 30 participants will be sought, though this will remain flexible. Interviews (above ten) will continue until a sufficient depth of understanding is achieved relating to the research objectives (Braun & Clarke 2021).

A varied sample will be sought, including a mixture of males and females, a range of ages (50 and over), and different occupational contexts (including type of workplace and roles held). Retirement is predicted to be the most reported transition, but recruitment will be open to other work-related transitions (such as role or career change).

### Recruitment

Purposive sampling will be utilised to ensure a broad sample is recruited, representing a range of transitions, participant characteristics and contexts. As this research aims to explore a range of work-related transitions, rather than just focussing on retirement, this sampling strategy is appropriate to ensure different perspectives, contexts and experiences are included (Robinson et al., 2013). A simple matrix will be created to ensure key groups are represented and a diverse selection is recruited (including sex, transition type etc).

Various recruitment techniques will be utilised to ensure a wide range of participants are identified. The study will be advertised via poster, leaflet or email in a selection of relevant ‘older adult’ locations, including community centres, retirement groups, mailing lists such as the University of York Pension groups, local cafes and businesses and communal spaces and on social media.

If required, for recruitment purposes, study information packs containing an invite letter, participant information sheet, and example consent form, may also be available in some of the relevant locations (described above).

### Consent process

Potential participants will be invited to contact the lead researcher (RW) by telephone or email to express their interest in taking part. The researcher will explain what the study involves and if the potential participant is interested, the researcher will post or email the participant the study information pack (including a consent form, participant details form, participant information sheet and invite letter) along with a stamped addressed envelope (if postal). The participant information sheet describes the study purpose and processes involved, including what would be involved for the participant, the withdrawal procedure, confidentiality and data management.

There will be several options for obtaining informed consent from participants: 1) the participant returns a competed and signed consent form in the post, 2) the participant returns a completed and signed consent form via email, or 3) the participant provides audio-recorded verbal consent prior to their qualitative interview. Once the consent form (and participant details form) is returned (via post or email), the researcher will make contact via phone or email (participant preference) to arrange the interview.

Participants will also complete a participant details form. These details are collected for contact and sampling purposes.

### Procedure (data collection)

The researcher will arrange a day and time to conduct the interview, arranged to be most convenient for the participant. Semi-structured individual interviews will take place over the telephone or using a remote online platform such as Zoom (dependant on participant preference) and will be audio-recorded. A predefined topic guide will be used to guide the interviews. The topic guide has been reviewed by PPI advisory group members with experience of undertaking a later-life transition and refined accordingly. A mock interview was also held with a PPI member to test the topic guide, consent and audio-recording processes. Initially the topic guide will be piloted with two participants and will be refined as needed for subsequent interviews. Interviews will be conducted by the lead researcher (RW) and will last approximately 30 to 45 minutes.

#### Example questions from the topic guide

1. *What was your experience of the transition?* PROMPT - how did you feel prior to [*retirement/other*], how did you find the transition period? Did you feel well supported (employees/family)?
2. *Can you describe any difficulties you experienced with your mental health and wellbeing?*
3. *What, if anything, do you think would have helped you during this time?*

### Analysis

The audio-recordings from the interviews will be transcribed verbatim by the lead researcher (RW). The transcripts will be initially read by the lead researcher, to allow the researcher to become familiar with the data and will then be descriptively coded. Following guidelines from Braun and Clarke (2022), a second researcher will not be sought (to code a selection of the transcripts) to test validity of the coding. Within reflective thematic analysis, data meanings are subjective, and codings are open to different interpretations. Attempting to use a second researcher to code transcripts to assess inter-rater reliability is therefore considered to be redundant as reflective thematic analysis is flexible and will vary across researchers. An appropriate coding software such as NVivo will be used.

The qualitative method of reflective thematic analysis will be used to inductively identify important or interesting patterns and themes within the data (Braun & Clarke, 2006). The framework put forward by Braun and Clarke (2006), describes a six-step process for conducting thematic analysis. This process will be followed for the analysis of the study data. Thematic analysis is an appropriate method of analysis to understand experiences, thoughts, and behaviours within a set of interview data (Braun & Clarke, 2012).

#### The six steps of thematic analysis are

##### Step one

Become familiar with the data – by reading and re-reading the transcripts and making notes.

##### Step two

Generate initial codes - meaningful organisation of the data in light of the study aims and research questions.

##### Step three

Search for themes - identifying interesting or significant patterns within the data to produce preliminary themes.

##### Step four

Review themes - modify and develop the themes identified in step three, considering their level of coherency and uniqueness from each other.

##### Step five

Define themes - confirming the themes and subthemes, how they interact and the meaning behind them.

##### Step six

The write-up - drafting of the full report to detail the study processes and findings.

### Data management

Participant identifiable materials (i.e., consent forms, participant details forms) will be stored on secure password-protected computers or in locked filing cabinets at the University of York and will only be accessible by the research team. Audio-recordings will be given a unique code and will be uploaded onto a secure password-protected computer. These recordings will be anonymised during the transcription process with any identifiable information removed. Audio-recordings will be deleted from the voice recorder once the transcription process is complete. Signed consent forms, audio-recordings and transcripts will be securely stored for up to 10 years after the study has completed, after which will be deleted or destroyed.

### Ethics and Risks

Ethical approval for this study was obtained from the University of York, Department of Health Sciences Research Governance Committee on 19^th^ May 2023 (ref. HSRGC/2023/566/B). The study data will be confidential and anonymised throughout the study period and dissemination. Participants will be allocated a unique ID for the study. The study is currently recruiting, and it is anticipated the study will be completed and written up by the end of 2025.

### Dissemination

This study will be written up as part of a PhD thesis and will be published in a relevant academic journal and may be presented at research conferences. All participants involved in the study will be offered a summary of the findings. This will be sent via email with a postal option for participants where email is not a feasible option.

## Discussion

This qualitative study will add to the limited literature in this area, to provide an in-depth insight into how work-related later life transitions are experienced by people who have experienced a negative impact on their mental health or wellbeing due to the transition. The study will help to inform future research looking to develop supports for people undertaking such transitions, by providing a richer understanding of the context, experience and support surrounding the different transitions.

One potential risk to this study is ensuring a diverse sample covering the different types of work-related transitions, the context of the transition (voluntary vs unvoluntary) and the different types of job roles. To mitigate this risk, the study will employ a range of recruitment methods to target potential participants across different geographical locations, organisations, and groups. The recruitment strategy will be monitored and refined if required.

In summary, the study will explore the experience of those who have recently undertaken a work-related later life transition, in relation to their mental health and wellbeing. This will help inform future research to develop suitable forms of support of people undertaking such transitions.

## Data Availability

Deidentified qualitative interview data will be held for 10 years after it is collected, and will be used for the purposes of the wider PHD study.

## Contribution of authors

As this study forms part of a PhD, the lead author (PhD student) has led on the conception, design and writing of this protocol draft. Co-authors (PhD supervisors) have contributed in supervision to the development of the protocol, and the reviewing and editing of this manuscript.

